# Optimal Control For A Crossover Cholera Mathematical Model Using Fractal (Variable-Fractional) Ψ-Caputo Derivative With Nonstandard Kernel

**DOI:** 10.1101/2025.03.31.25324982

**Authors:** Seham M. AL-Mekhlafi, Ebenezer Bonyah

## Abstract

This paper introduces an optimal control strategy for cholera’s crossover mathematical model. The proposed model integrates Ψ-Caputo fractal variable-order derivatives, fractal fractional-order derivatives, and integer-order derivatives across three distinct time intervals, utilizing a simple non-standard kernel function Ψ(*t*). A comprehensive stability analysis of the model’s steady states is conducted. The model’s results are compared with real-world data from the cholera outbreak in Yemen. Following this, an optimal control problem is formulated within the crossover framework. To numerically solve the resulting optimality system, a discretized non-standard -finite difference method is developed. Numerical simulations and comparative studies are presented to demonstrate the method’s applicability and the efficiency of the approximation approach. The key finding of this study highlights that the crossover-controlled system proves to be the most effective approach for mitigating and controlling the spread of cholera.

## 1 Introduction

Cholera, an acute diarrheal disease caused by ingesting food or water contaminated with Vibrio cholerae, remains a significant global health challenge [1]. Despite advancements in healthcare and sanitation, periodic outbreaks continue to pose threats, particularly in regions with limited access to clean water and adequate medical resources [2]. Mathematical modeling of cholera transmission dynamics has proven to be a valuable tool for understanding the spread of the disease and designing effective control strategies [3, 4, 6].

Classical cholera models typically employ integer-order derivatives to describe the dynamics of the disease. However, such models may not adequately capture the complexity and memory effects inherent in biological and epidemiological systems. Recent advancements in fractional calculus have opened new avenues for modeling these dynamics with greater accuracy. Fractional derivatives, which extend the concept of integer-order differentiation to non-integer orders, are particularly effective in incorporating memory effects and non-local interactions [24, 18]. In this context, the Ψ-Caputo derivative, a generalized form of the Caputo fractional derivative, offers a flexible framework for incorporating variable-order fractional dynamics. This allows for modeling systems where the order of differentiation varies with time or other parameters, providing a more realistic representation of complex systems [19, 21, 22].

Fractal fractional calculus has emerged as a transformative approach in mathematical epidemiology, providing new insights into the dynamics of infectious diseases like cholera. Unlike classical and standard fractional derivatives, fractal fractional derivatives combine the concepts of fractional-order differentiation and fractal geometry, enabling the modeling of systems with complex, irregular, and memory-dependent behaviors. In the context of cholera, such models are particularly effective in capturing the heterogeneous nature of disease transmission, influenced by spatial variability, environmental factors, and human behavior. The incorporation of fractal dynamics allows for the representation of transmission pathways that are not only dependent on time but also exhibit fractal-like properties, such as the clustering of cases in contaminated water sources or the irregular spatial spread of outbreaks [23].

The application of optimal control theory to fractional-order models is a burgeoning area of research. Optimal control seeks to determine the best strategies to influence system dynamics, often subject to constraints and objectives. When applied to cholera models, it facilitates the design of interventions that minimize disease prevalence and associated costs while optimizing resource allocation[4, 5, 20]. Integrating variable-order fractional derivatives into optimal control frameworks enhances their ability to capture the intricacies of real-world systems and offers a promising approach to public health decision-making.

This study focuses on the development of an optimal control strategy for cholera’s crossover model based on (variable-fractional) fractal order Ψ-Caputo derivative. The crossover model incorporates elements that describe both direct and indirect transmission dynamics, as well as intervention measures such as vaccination, water treatment, and public health campaigns. By leveraging the flexibility of variable-fractional derivatives and the rigor of optimal control theory, the proposed approach aims to address the limitations of classical models and provide actionable insights for cholera management.

The remainder of this work is organized as follows: Section 2 A review of essential concepts and results from the literature is presented. Section 3 introduces the foundational model and provides its mathematical analysis. Section 4 presents the formulation of the crossover cholera model and the Ψ-Caputo derivative framework and introduces the optimal control problem, including the objective function and constraints for the crossover model. Section 5 discusses the numerical methods employed for simulation and analysis. Section 6 presents numerical simulations that illustrate the performance and accuracy of the crossover optimization system. Finally, Section 7 provides conclusions and recommendations for future research.

## 2 Preliminary Information and Notation

In this section, we recall some important definitions of the fractional calculus used throughout the remaining sections of this paper.

### Proposition 2.1.

*[24] Let* 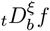 *and* 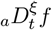 *represent the right-sided and left-sided Riemann-Liouville (RL) fractional derivatives of f* (*t*), *respectively. Similarly, let* 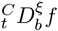 *and* 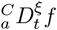 *denote the right-sided and left-sided Caputo fractional derivatives of f* (*t*). *Here, ξ* ∉ ℕ, *and for n* = [ℜ(*ξ*)] + 1,

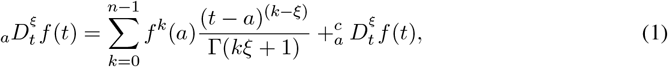

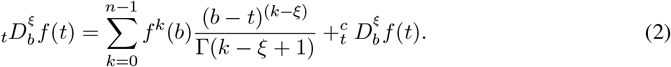

### Proposition 2.2.

*Let f* (*t*) *be a sufficiently differentiable function on* [*a, b*].

1. *If*

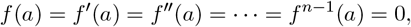

*then the left-sided Riemann-Liouville fractional derivative of f* (*t*) *is equal to the left-sided Caputo fractional derivative of f* (*t*), *i*.*e*.,

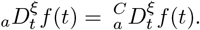 *This result holds because the initial conditions ensure that the additional terms in the Caputo fractional derivative formulation vanish*.
2. *Similarly, if*

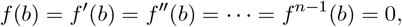

*then the right-sided Riemann-Liouville fractional derivative of f* (*t*) *is equal to the right-sided Caputo fractional derivative of f* (*t*), *i*.*e*.,

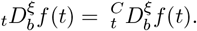

### Definition 2.1.

*Assuming f* : [*a, b*] → ℝ, *be integrate*, 0 < *ξ, and* Ψ ∈ *C*^1^([*a, b*]) *an increasing function* ∀ *t* ∈ [*a, b*] *such that* Ψ′≠ 0, *with order ξ, the definition of the fractional integral of* Ψ*-Riemann-Liouville for f is [7, 8]:*

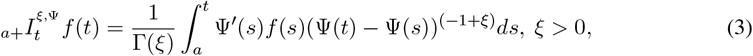

Γ(*ξ*) *represents the function of Gamma. The* Ψ(*t*) = *Ln*(*t*), *and* Ψ(*t*) = *t, equations (3) are equivalent to the Hadamard fractional integral and the Riemann-Liouville problem, respectively*.

### Definition 2.2.

*Suppose that n* ∈ ℕ *and* Ψ, *f* ∈ *C*^*n*^([*a, b*], ℝ) *be two functions; for every t* ∈ [*a, b*], Ψ *is growing and* Ψ′ ≠ 0. *The* Ψ*-Riemann-Liouville fractional derivative of order ξ for f is defined as [7, 8]:*

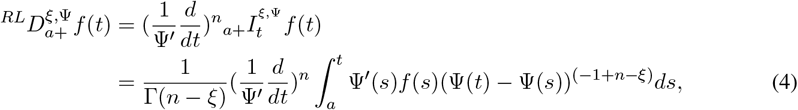

*where n* = [*ξ*] + 1.

### Definition 2.3.

∀*t* ∈ [*a, b*], *let theref*, Ψ ∈ *C*^*n*^([*a, b*], ℝ) *be two functions such that* Ψ *is rising, and* 0 ≤ Ψ′. *[9] defines the fractional derivative of* Ψ*-Caputo with order ξ*.

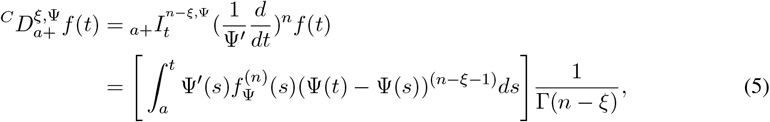

*where* 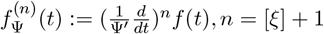.

This work will expand the fractional derivative of Ψ-Caputo [9] to fractal fractional order and variable order fractal Ψ-Caputo derivative, as follows:

First, we can use the following definition of the fractal fractional derivative of Ψ-Caputo [10].

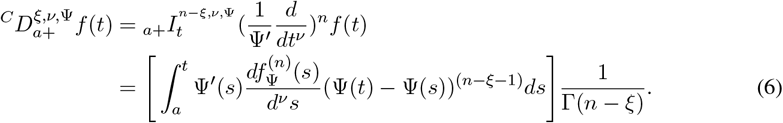

Additionally, the following is the Ψ-Caputo fractional derivative for the fractal variable order. [10].

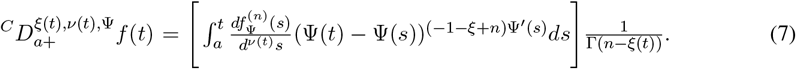

According to (6) and (7), the well-known Caputo fractal fractional and variable order fractal derivatives are obtained if Ψ(*t*) = *t*. Furthermore, the Caputo-Katugampola derivative fractal fractional and variable order fractal derivatives are obtained if Ψ(*t*) = *t*^*ϵ*^, *ϵ* ≥ 0.

## 3 The Basic Model

In this section. The basic model of cholera disease, which is given in [4], is introduced. The following fundamental populations are included in this model: Susceptible 𝕊, infectious 𝕀, quarantined ℚ, and recovered ℝ are the different categories of humans. The population of free bacteria in the environment, 𝔹, is then taken into consideration as well. This is a crucial requirement since a healthy person must consume environmental bacteria to get infected, which eliminates the bacteria from the aquatic medium. All parameters in the model are defined in Table 1. Then the basic model is given as follows:

**Table 1:** Key Parameters and Initial Conditions Used in the Model [4].

| Parameter | Modified Description | Value |
| --- | --- | --- |
| $\Lambda$ | Population recruitment rate | $28.4N(0)/365,000$ (person day<sup>-1</sup>) |
| $\mu$ | Rate of natural mortality | $1.6 \times 10^{-5}$ (day<sup>-1</sup>) |
| $\beta$ | Consumption rate | 0.01891 (day<sup>-1</sup>) |
| $\kappa$ | Saturation threshold | $10^7$ (cell/ml) |
| $\omega$ | Decline rate of immunity | $0.4/365$ (day<sup>-1</sup>) |
| $\delta$ | Isolation rate | 1.15 (day<sup>-1</sup>) |
| $\epsilon$ | Recovery probability rate | 0.2 (day<sup>-1</sup>) |
| $\lambda_1$ | Fatality rate (infected) | $6 \times 10^{-6}$ (day<sup>-1</sup>) |
| $\lambda_2$ | Mortality rate (quarantined) | $3 \times 10^{-6}$ (day<sup>-1</sup>) |
| $\eta$ | Pathogen release rate (infected) | 10 (cell/ml day<sup>-1</sup> person<sup>-1</sup>) |
| $d$ | Pathogen degradation rate | 0.33 (day<sup>-1</sup>) |
| $\rho$ | Transmission coefficient | 0.01891 (cell/ml day<sup>-1</sup> person<sup>-1</sup>) |
| $S(0) = S_0$ | Initial number of susceptible individuals | 28,249,670 (person) |
| $I(0) = I_0$ | Initial number of infected individuals | 750 (person) |
| $Q(0) = Q_0$ | Initial number of quarantined individuals | 0 (person) |
| $R(0) = R_0$ | Initial number of recovered individuals | 0 (person) |
| $B(0) = B_0$ | Initial bacterial concentration | $275 \times 10^3$ (cell/ml) |

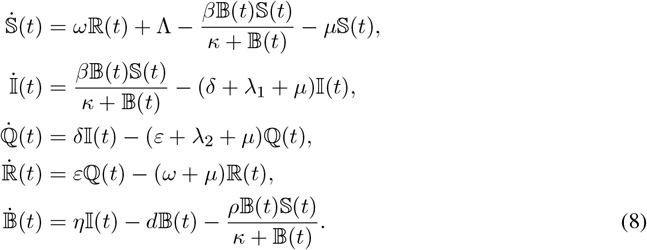

### 3.1 Analysis of the Model

In this study, we assume that system (1)’s beginning conditions are nonnegative:

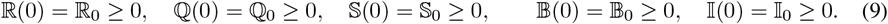

### 3.2 Positivity and boundedness of solutions

Our first lemma demonstrates the biological significance of the models (8)–(9) under consideration.

#### Lemma 1

For all *t* ≥ 0, the solutions (𝕊(*t*), 𝕀(*t*), ℚ(*t*), ℝ(*t*), 𝔹(*t*)) of system (8) are nonnegative with nonnegative initial conditions (9) in 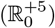.

**proof**: We have

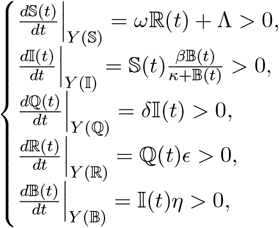

where *Y* (ν) = *α*(t) = 0, *α* = {ℚ, 𝕀, 𝕊, ℝ, 𝔹}. Therefore, any solution of system (8) is such that (𝕊(*t*), 𝕀(*t*), ℚ(*t*), ℝ(*t*), 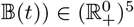 for all *t* ≥ 0.

Next Lemma 2 shows that it is sufficient to consider the dynamics of the flow generated by (8)–(9) in a certain region Ω.

#### Lemma 2

Let

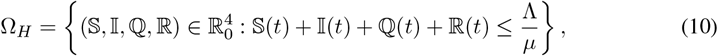

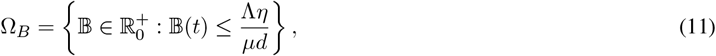

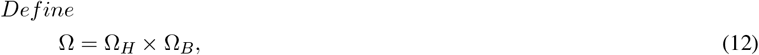

If 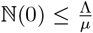 and 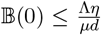, then the region Ω is positively invariant for model (8) with nonnegative initial conditions (9) in 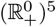.

**Proof**: Let’s divide system (8) into two components: the pathogen population, or 𝔹(*t*), and the population of human 𝕀(*t*), 𝕊(*t*), ℝ(*t*), and ℚ(*t*). The first four equations of system (8) are added to give *N*.

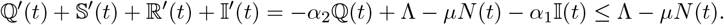

Given the assumption that 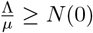, we deduce that 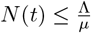. It follows that

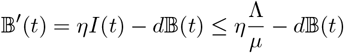

for the population of pathogens. If 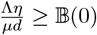, then 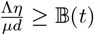.

We can infer from (10) and (11) that for all *t* ≥ 0, *B*(*t*) and *N* (*t*) are bounded. As a result, each system (8) solution with an initial condition in Ω.

### 3.3 Local stability analysis of the disease-free equilibrium

To analyze the local asymptotic stability of the disease-free equilibrium (DFE) of the given system, we follow these steps:

Step 1: Identify the Disease-Free Equilibrium (DFE)

The disease-free equilibrium occurs when the infection variables are zero.

At equilibrium, the system reduces to the case where only the susceptible and other healthy compartments remain. Define the DFE as:

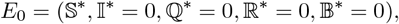

where 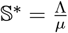 satisfies the non-disease dynamics.

Step 2: Compute the Jacobian Matrix

The local stability of *E*_0_ is determined by computing the Jacobian matrix *J* of the system at *E*_0_. The Jacobian is obtained by linearizing the system around the equilibrium.

Define the state vector:

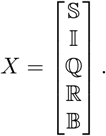

The Jacobian matrix *J* consists of the partial derivatives:

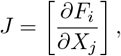

where *F*_*i*_ are the right-hand side functions of the system.

Step 3: Evaluate the Key Submatrix for Infection Dynamics

Since the disease-free stability depends primarily on the infected compartments (*I* and *B*), we focus on the relevant submatrix involving the infection-related terms:

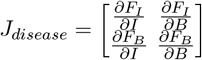

From the given system, the key equations governing infection dynamics are:

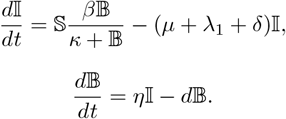

Linearizing around *E*_0_, we approximate:

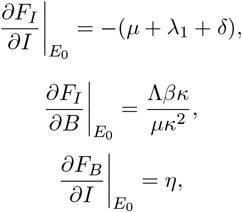

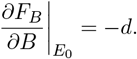

Thus, the infection submatrix is:

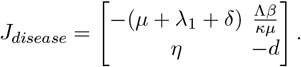

Step 4: Compute the Basic Reproduction Number *R*_0_ [26]

The threshold parameter that determines stability is the spectral radius (dominant eigenvalue) of the next-generation matrix [27]. The basic reproduction number is given by:

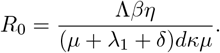

Step 5: Stability Analysis

- If *R*_0_ < 1, all eigenvalues of *J*_*disease*_ have negative real parts, implying local asymptotic stability of the disease-free equilibrium.
- If *R*_0_ > 1, there exists a positive eigenvalue, implying the instability of the disease-free equilibrium.

**Theorem:** The Cholera-free equilibrium point *E*_0_ is locally asymptotically stable if *R*_0_ < 1 and unstable if *R*_0_ > 1.

*Proof*. To analyze the local stability of the system (8) at an equilibrium point, we need to compute the Jacobian matrix and evaluate its eigenvalues at the given equilibrium.

Step 1: Compute the Jacobian Matrix The system of equations (8) is:

The Jacobian matrix at disease free Equlibrum *J*(*E*_0_) is the matrix of first-order partial derivatives at *E*_0_ is given as follows:

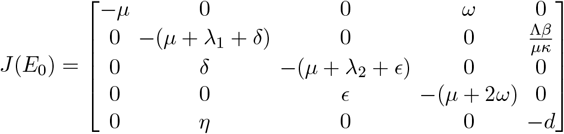

Step 2: Compute the characteristic polynomial *det*(*J*(*E*_0_) − *σI*) = 0.

It is easy to see that the first negative eigenvalue is *σ*_1_ = −*µ* The remaining eigenvalues are obtained by giving the characteristic polynomial of the following matrix

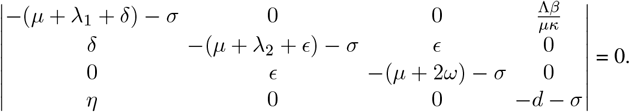

The first eigenvalue is clearly −*µ*, which is negative. The block related to *Q* and *R* involves the submatrix:

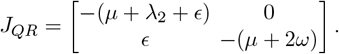

To compute *det*(*J*_*QR*_ − *σI*) = 0, since all entries are negative, the eigenvalues of this block are negative. The infection-related submatrix is:

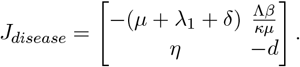

The determinant of this matrix is:

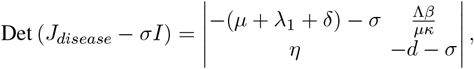

we have the following characteristic equation:

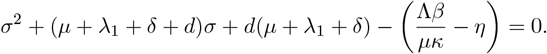

Now, using the given expression for *R*_0_:

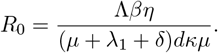

We rewrite the quadratic equation as:

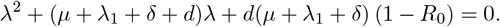

**To applying the Routh-Hurwitz criterion [28] to the quadratic equation:**

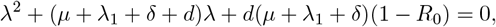

we define the standard quadratic form:

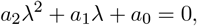

where:

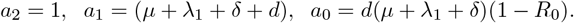

Routh-Hurwitz Stability Conditions for a Quadratic Equation

For the system to be stable, all roots of the characteristic equation must have negative real parts. The necessary and sufficient conditions for this are:

1. *a*_1_ > 0,
2. *a*_0_ > 0.

Applying the Conditions

- First Condition: *a*_1_ > 0 Since *µ, λ*_1_, *δ*, and *d* are all positive parameters, we have:

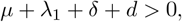

which is always true.
- Second Condition: *a*_0_ > 0,

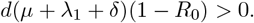 Since *d* > 0 and *µ* + *λ*_1_ + *δ* > 0, the sign of *a*_0_ depends on (1 − *R*_0_).
- If *R*_0_ < 1, then (1 − *R*_0_) > 0 and *a*_0_ > 0, ensuring stability.
- If *R*_0_ > 1, then (1 − *R*_0_) < 0 and *a*_0_ < 0, leading to instability (one root will be positive).
- If *R*_0_ = 1, then *a*_0_ = 0, meaning one root is at zero, indicating a critical stability condition.

### 3.4 Global stability analysis of the disease-free equilibrium

To extend our analysis from local stability to global stability, we need to show that the disease-free equilibrium *E*_0_ is globally asymptotically stable when *R*_0_ < 1. This means that all solutions of the system converge to *E*_0_ as *t* → ∞, regardless of initial conditions.

Step 1: Construct a Lyapunov Function

A common approach to proving global stability is by constructing a Lyapunov function, which decreases over time and satisfies certain conditions.

Define the Lyapunov function:

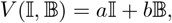

where *a* and *b* are positive constants to be determined. Taking the time derivative along the system trajectories:

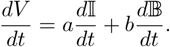

Substituting the equations for 𝕀 and 𝔹:

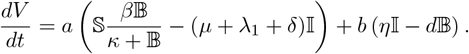

Evaluating at the disease-free equilibrium where 𝕊* is constant:

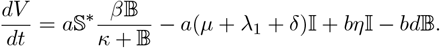

Rewriting:

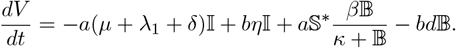

To ensure 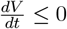, choose *a* and *b* such that the term −*a*(*µ* + *λ*_1_ + *δ*)𝕀 + *bη*𝕀 is negative. Setting:

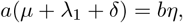

which implies:

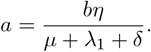

Substituting:

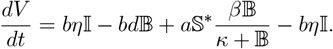

Factoring:

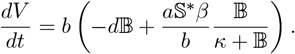

Since we defined:

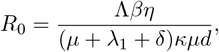

we rewrite:

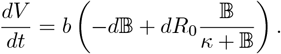

If *R*_0_ < 1, then:

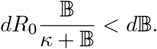

Thus, 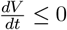, and *V* (𝕀, 𝔹) is a Lyapunov function, ensuring that (𝕀, 𝔹) → (0, 0) globally.

Step 2: LaSalle’s Invariance Principle

By LaSalle’s Invariance Principle, all solutions of the system approach the largest invariant set where 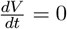, which is only at the DFE when 𝕀 = 0, and 𝔹 = 0.

Therefore, for *R*_0_ < 1, the DFE is globally asymptotically stable.

### 3.5 Stability analysis of the endemic equilibrium

When *R*_0_ > 1, the system allows for an endemic equilibrium where the disease persists in the population. We now analyze its existence and stability.

Step 1: Find the Endemic Equilibrium: The endemic equilibrium *E*^*^ is a steady-state solution where 𝕀*≠ 0 and 𝔹*≠ 0. Denoting the equilibrium values as:

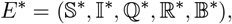

we solve the steady-state equations.

From the bacteria equation:

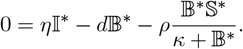

Solving for 𝔹*:

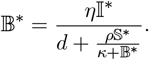

Solve for 𝕀*: From the infected equation:

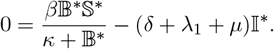

Rearrange for 𝕀*:

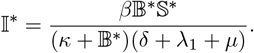

Solve for 𝕊*: From the **susceptible equation**:

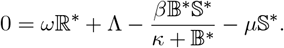

Rearrange for 𝕊*:

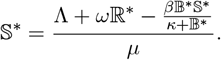

Solve for ℚ* and ℝ*: From the quarantined equation:

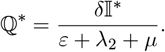

From the recovered equation:

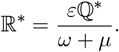

### 3.6 Local stability analysis of the endemic equilibrium

To prove the local stability of the endemic equilibrium, we analyze the Jacobian matrix of the system at the endemic equilibrium and check the eigenvalues. The endemic equilibrium is locally asymptotically stable if all eigenvalues have negative real parts.

Step 1: Compute the Jacobian Matrix at *E*^*^:

The Jacobian matrix *J* is givn by:

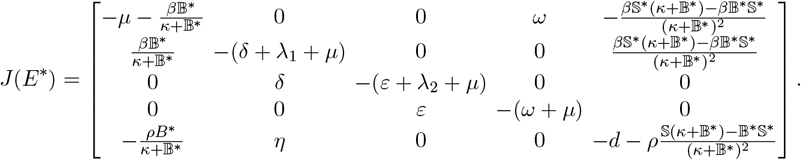

Step 2: Compute Eigenvalues To check local stability, we solve:

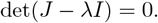

Expanding this determinant is complex. Instead, we use the trace and determinant to infer stability.

Step 3: compute trace and Determinant conditions Trace of *J* is given by:

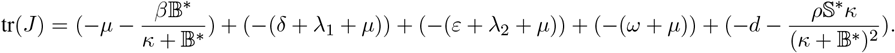

If tr(*J*) < 0, then the sum of eigenvalues is negative.

Also, if det(*J*) > 0, then all eigenvalues are negative or complex with negative real parts.

Using Routh-Hurwitz criteria, we check that all eigenvalues have negative real parts, proving local stability.

### 3.7 Global stability analysis of the endemic equilibrium

Global Stability Analysis of the Endemic Equilibrium To establish the global stability of the endemic equilibrium (𝕊*, 𝕀*, ℚ*, ℝ*, 𝔹*), we use Lyapunov’s method and LaSalle’s Invariance Principle.

Step 1: Define the Lyapunov Function A Lyapunov function is a scalar function *V* (𝕊, 𝕀, ℚ, ℝ, 𝔹) that is positive definite and decreasing along the trajectories of the system.

We define a Lyapunov function for the system:

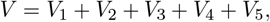

where each component is:

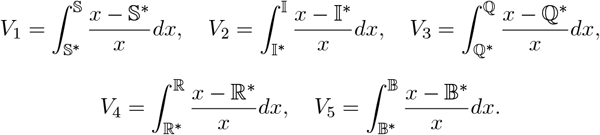

These functions satisfy:

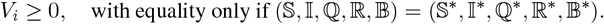

Step 2: Compute the Time Derivative 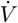

To establish global stability, we differentiate *V* along the system’s trajectories:

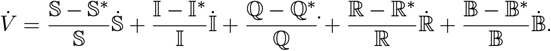

Substituting the system equations:

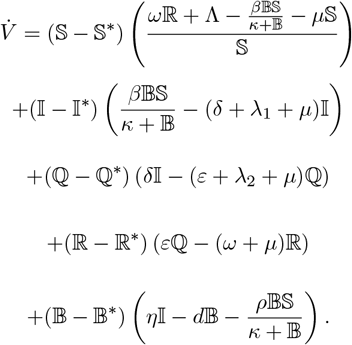

After simplification, we express 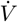 in the form:

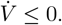

If 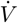 is negative semi-definite, we proceed with LaSalle’s Invariance Principle.

Step 3: Apply LaSalle’s Invariance Principle

- The largest invariant set where 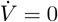 corresponds to the endemic equilibrium.
- According to LaSalle’s Invariance Principle, all trajectories approach the endemic equilibrium.

Thus, the endemic equilibrium (𝕊*, 𝕀*, ℚ*, ℝ*, 𝔹*) is globally asymptotically stable when *R*_0_ > 1.

## 4 Formulated Optimal Control Crossover Mathematical Model

In this section, we expanded the model of cholera [4] to the crossover optimal control cholera model defined in three-time intervals.

The spread of cholera is associated with a lack of access to sanitary facilities and clean water. To enhance water quality and prevent cholera outbreaks, one potential tactic is to provide chlorine water tablets (CWT) for water filtration.

We add a control function 𝕌(.) to the following crossover model, which denotes the percentage of vulnerable people with the ability to purify water using CWT (see [11]).

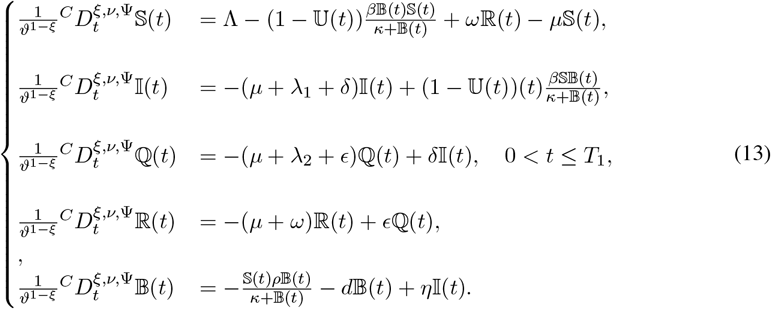

with initial conditions

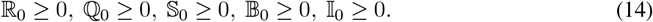

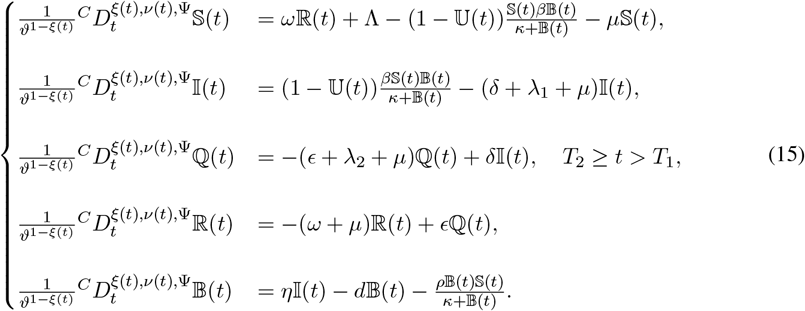

with initial conditions

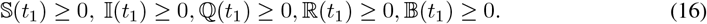

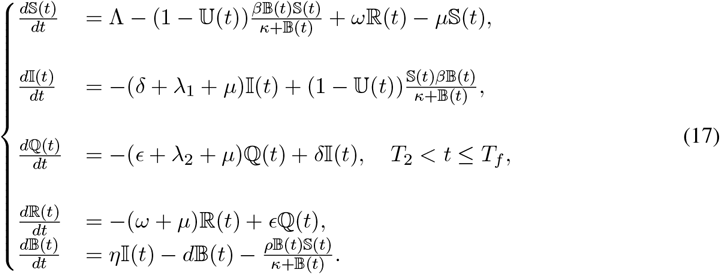

with

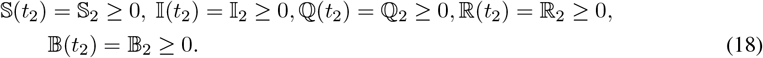

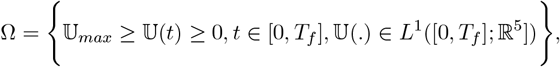

where 0 ≤ 𝕌_*max*_ ≤ 1.

The following is a definition of the cost functional:

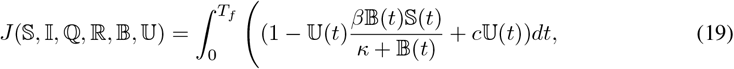

where *c* is a weight coefficient. Now, we want the objective to be to minimize the functional cost (19).

### 4.1 Necessary Optimality Conditions

The Hamiltonian function of the form

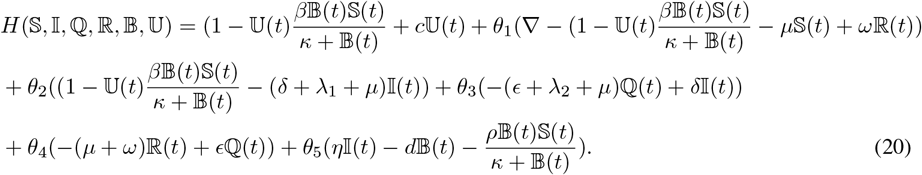

As in [12, 13, 14], Pontryagin’s maximum principle is used to determine the essential conditions.

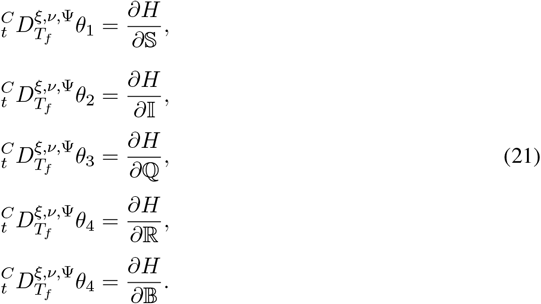

It is also necessary that

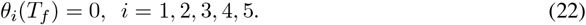

where the Lagrange multipliers are *θ*_*i*_, and

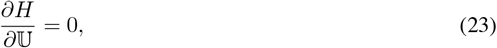

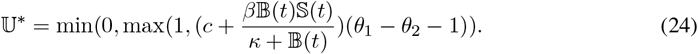

then we substitute (24) in the basic fractal fractional system, we have the following state system:

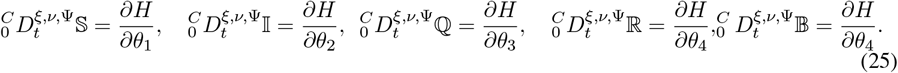

### 4.2 Optimal Control Crossover Problem: Descretization

With the corresponding state, let 𝕌* be the optimal control. Then 𝕊, 𝕀, ℚ, ℝ, 𝔹. The following is satisfied by the adjoint variables *θ*_*j*_ *j* = 1, 2, 3, 4, 5.

The adjoint equation in 0 < *t* ≤ *t*_1_ :

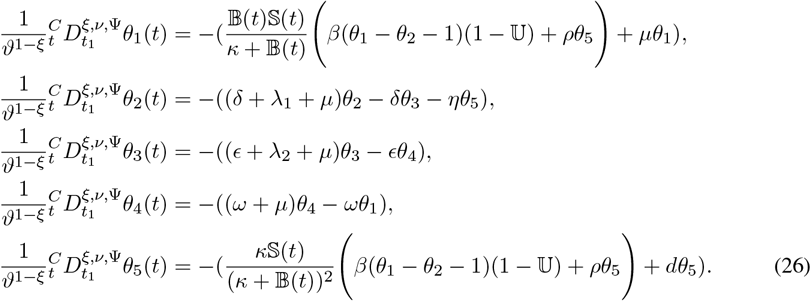

with transversality conditions

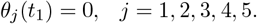

The adjoint equation in *t*_1_ < *t* ≤ *t*_2_ :

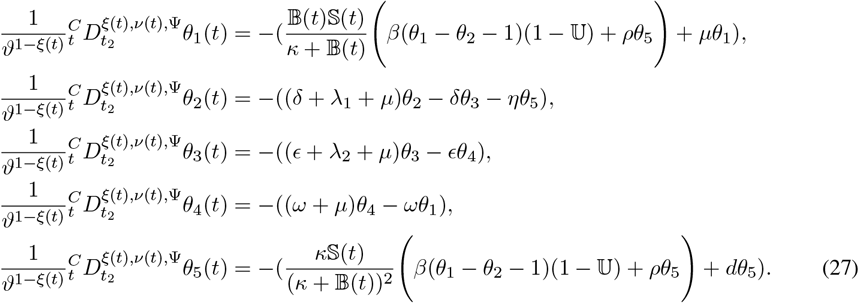

with transversality conditions

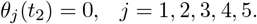

The adjoint equation in *t*_2_ < *t* ≤ *T*_*f*_ :

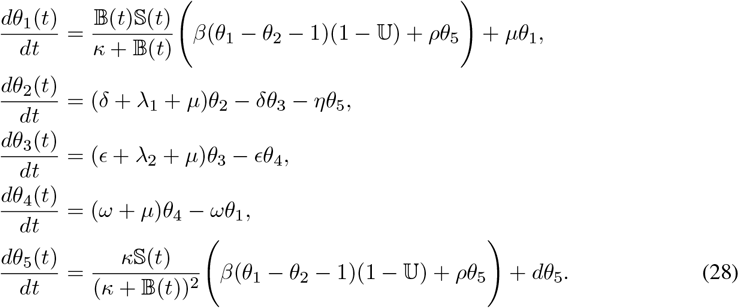

with transversality conditions

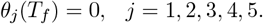

for practically every t in [0, *T*]. Furthermore, the control law is distinguished by

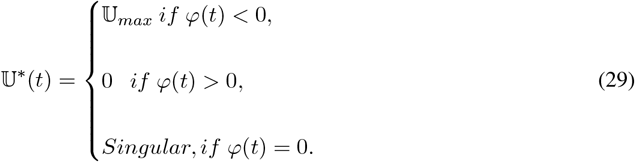

and

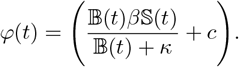

The state equations are now as follows after replacing 𝕌*(*t*) in (13), (15), and (17):

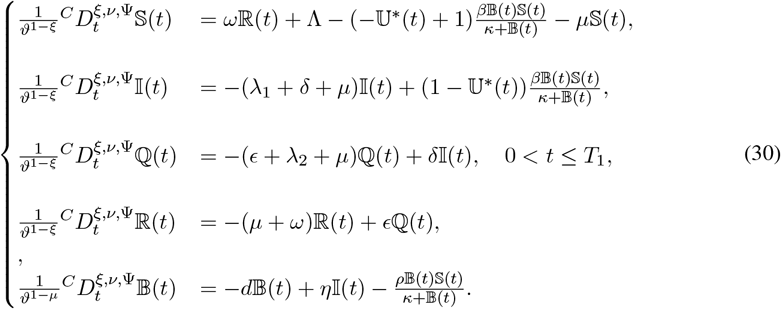

with initial conditions

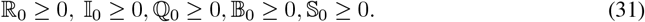

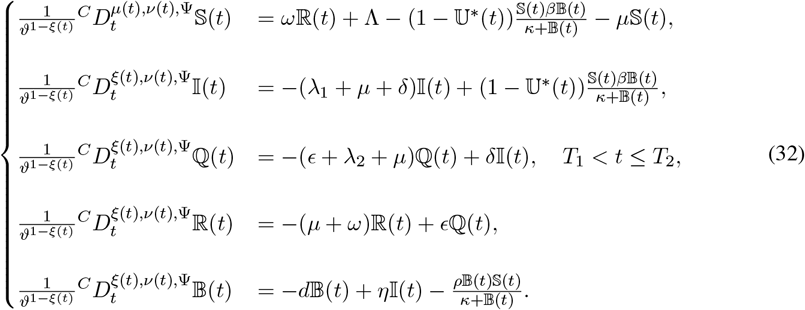

with initial conditions

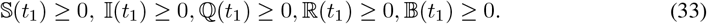

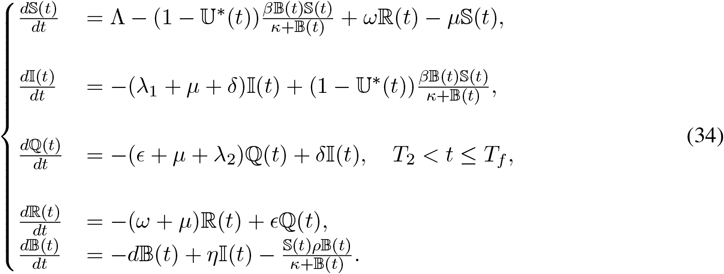

with

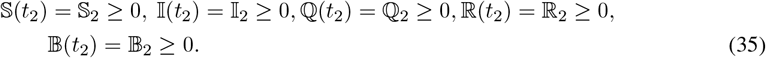

## 5 Numerical Approaches to the Suggested Models

### 5.1 Ψ-NSFDM

In this part, we introduce numerical techniques for solving (30-34) numerically. Examine the general form equation of the derivative for a crossover model that incorporates fractal fractional, fractal variable, and integer orders.

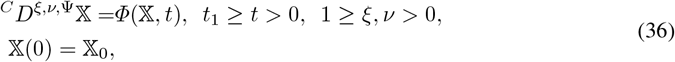

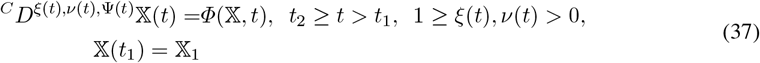

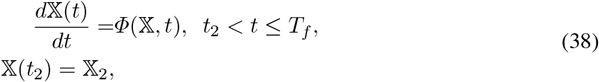

The following displays the discretization of (5) using a consistent mesh *h* = *t*_*j*+1_ − *t*_*j*_, *j* = 0, 1,, *N* − 1 and insert *f* (*t*) = 𝕏(*t*) and Ψ(*t*) = (𝔼*t* + 𝔻)^ς^, Also, by using ϖ(*h*) a positive function 1 ≤ ϖ(*h*) > 0, [15], and ς, 𝔻, 𝔼 are constants. 𝕏(*t*), is approximated using a non-standard method. Considering that [16, 17]

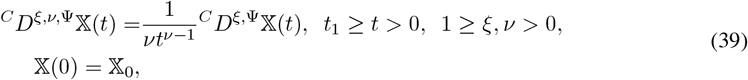

and

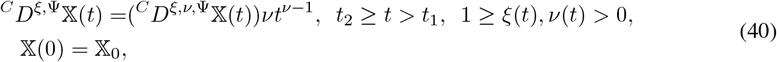

Also,

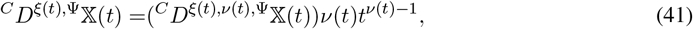

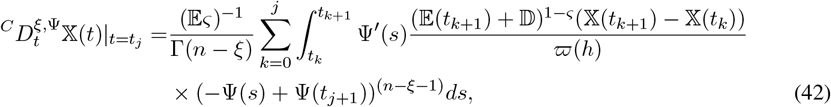

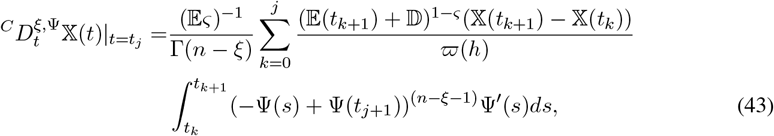

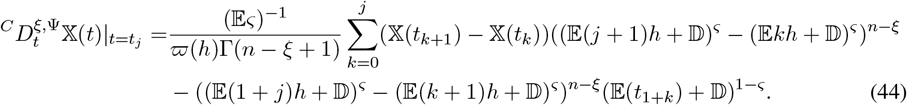

Also,

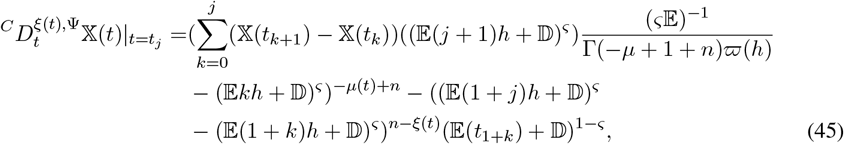

and,

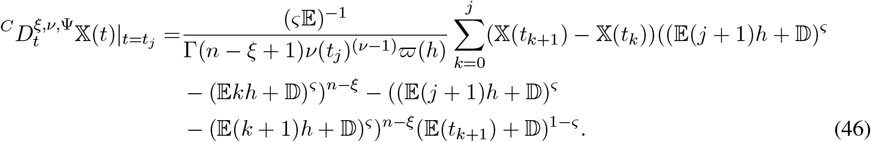

By employing the nonstandard finite difference method to solve (36) using (40) and (46), we have

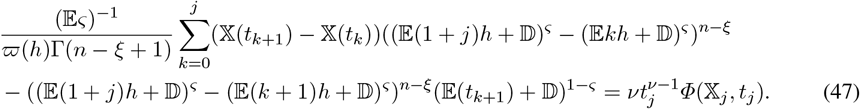

We have

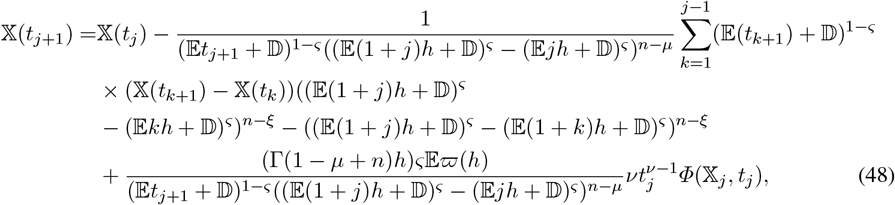

To solve (37) using (41) as follows:

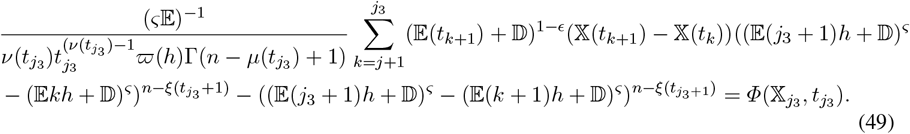

We have

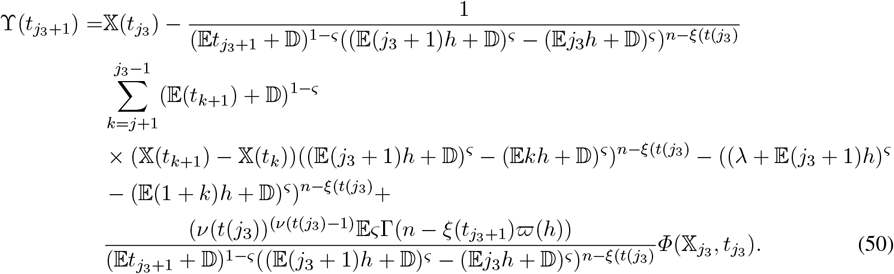

To solve (38) using NSFDM as follows:

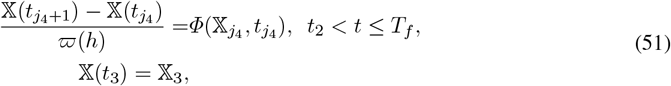

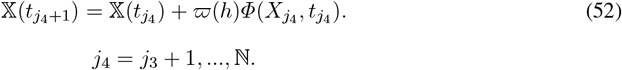

We discretize (26)-(28) with transitivity criteria using backward-in-time discretization, which yields an algebraic system of easily solved equations.

## 6 Discussion and Numerical Simulations

This section focuses on validating the experimental results and the analytical expressions derived in the previous parts. Real-world data on the number of cholera-infected individuals in Yemen, recorded between April 27, 2017, and April 15, 2018 [25], is utilized for this analysis. The numerical simulations are conducted based on the parameters and initial conditions outlined inside Table 1.

All calculations were performed with MATLAB 2021b on a computer equipped with an Intel(R) Core i7-3110M processor (1.80 GHz) and 8 GB of RAM.

Figure 1 shows the bifurcation diagram of *R*_0_ (the basic reproduction number) with respect to *β* (the transmission rate of cholera), where the blue line (*R*_0_ vs *β*) shows a linear relationship between *R*_0_ and *β*. As *β* increases, *R*_0_ increases proportionally, meaning higher transmission leads to a larger outbreak. Also, the red dashed line (*R*_0_ = 1 Threshold) represents a bifurcation threshold, where, When *R*_0_ = 1, the system transitions from a disease-free equilibrium to an endemic state where cholera persists. Below this line, the disease is controlled. Above this line, an epidemic occurs.

**Figure 1:**
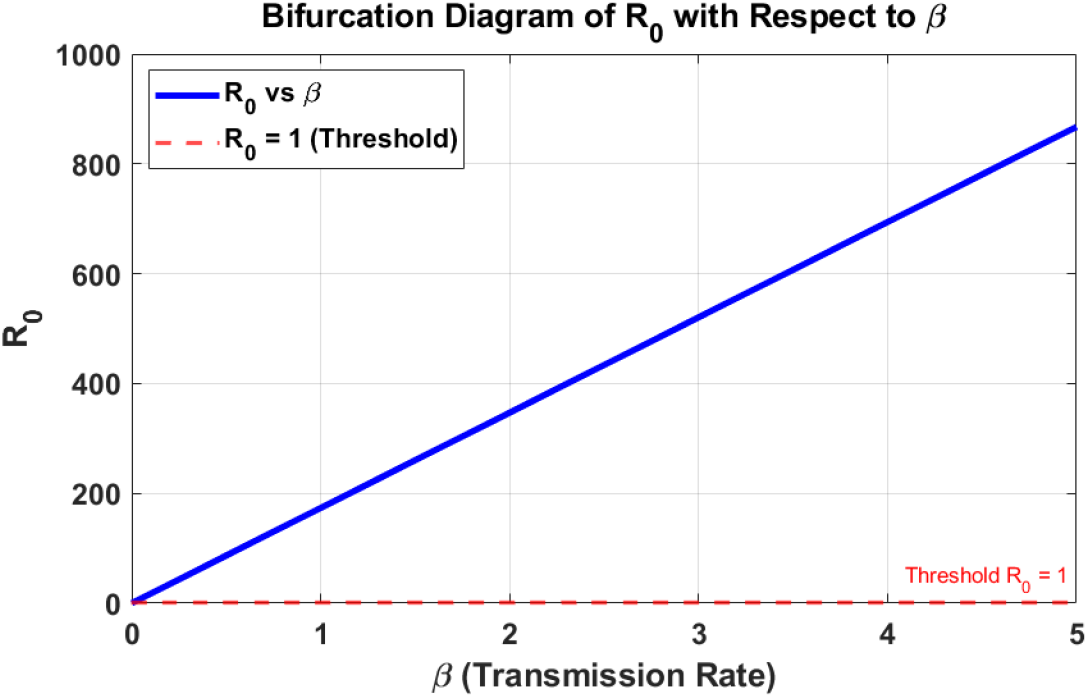
Bifurcation Diagram of *R*_0_ with respect to *β*

The bifurcation diagram in Figure 2 illustrates the relationship between the transmission rate (*β*) and the equilibrium number of infected individuals (*I*). Initially, at low *β*, the infection level is low, indicating that the disease struggles to spread. As *β* increases, *I* rises sharply and stabilizes at a high endemic level. The blue curve represents stable equilibria, where the system naturally settles, while the red points indicate unstable equilibria, suggesting a critical transition. This behavior implies that beyond a threshold *β*, the disease becomes persistent in the population. Effective control measures to reduce *β* are crucial for preventing endemic outbreaks and lowering infection levels.

**Figure 2:**
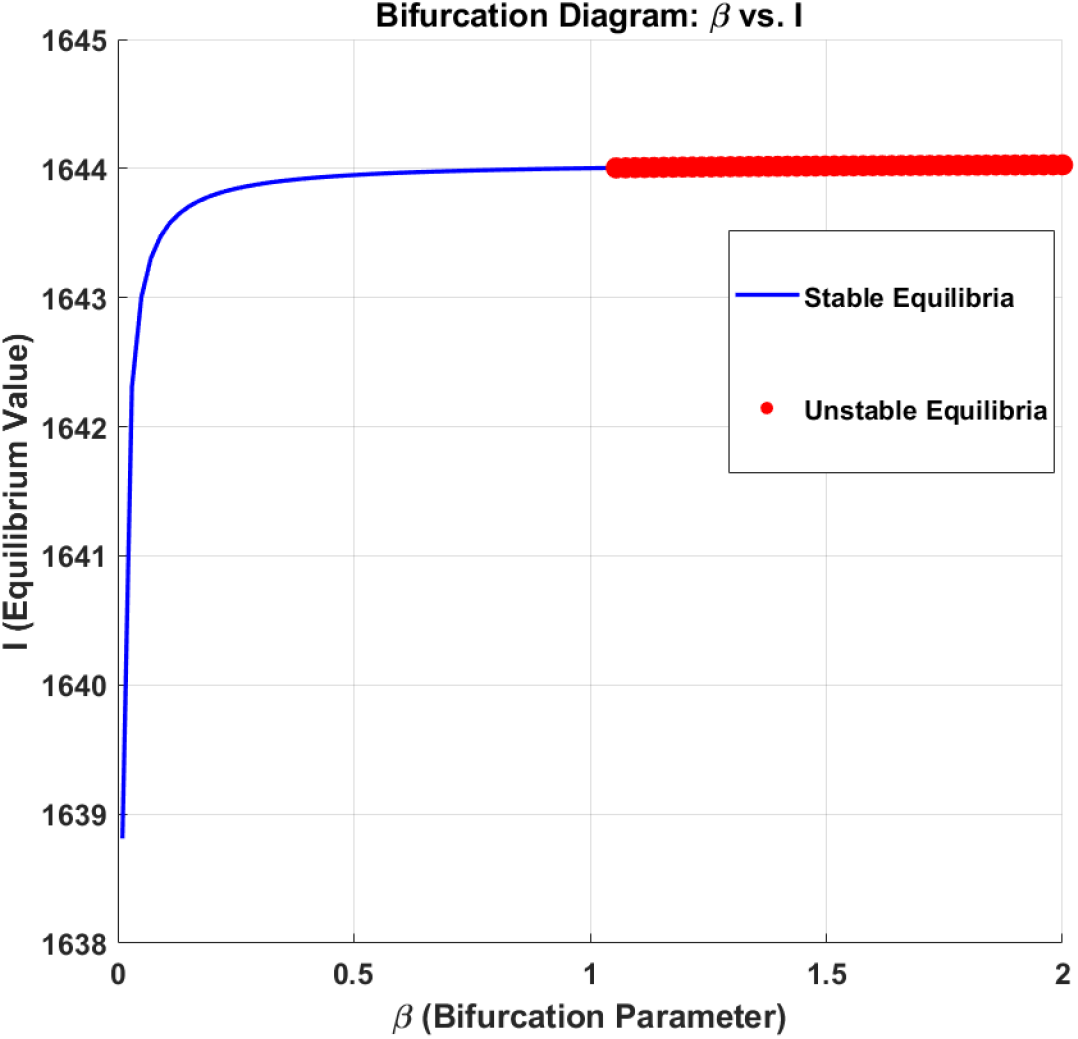
Bifurcation Diagram of *I* with respect to *β*

Figure 3 showcases the numerical simulation results for the piecewise system of cholera dynamics. It compares the real data (blue circles) with the approximate Solution (red line). The red curve, representing the approximate solution, closely follows the blue points, suggesting that the model provides a good fit to the real data. This indicates the model’s reliability in capturing the dynamics of cholera. Also, the temporal behavior

**Figure 3:**
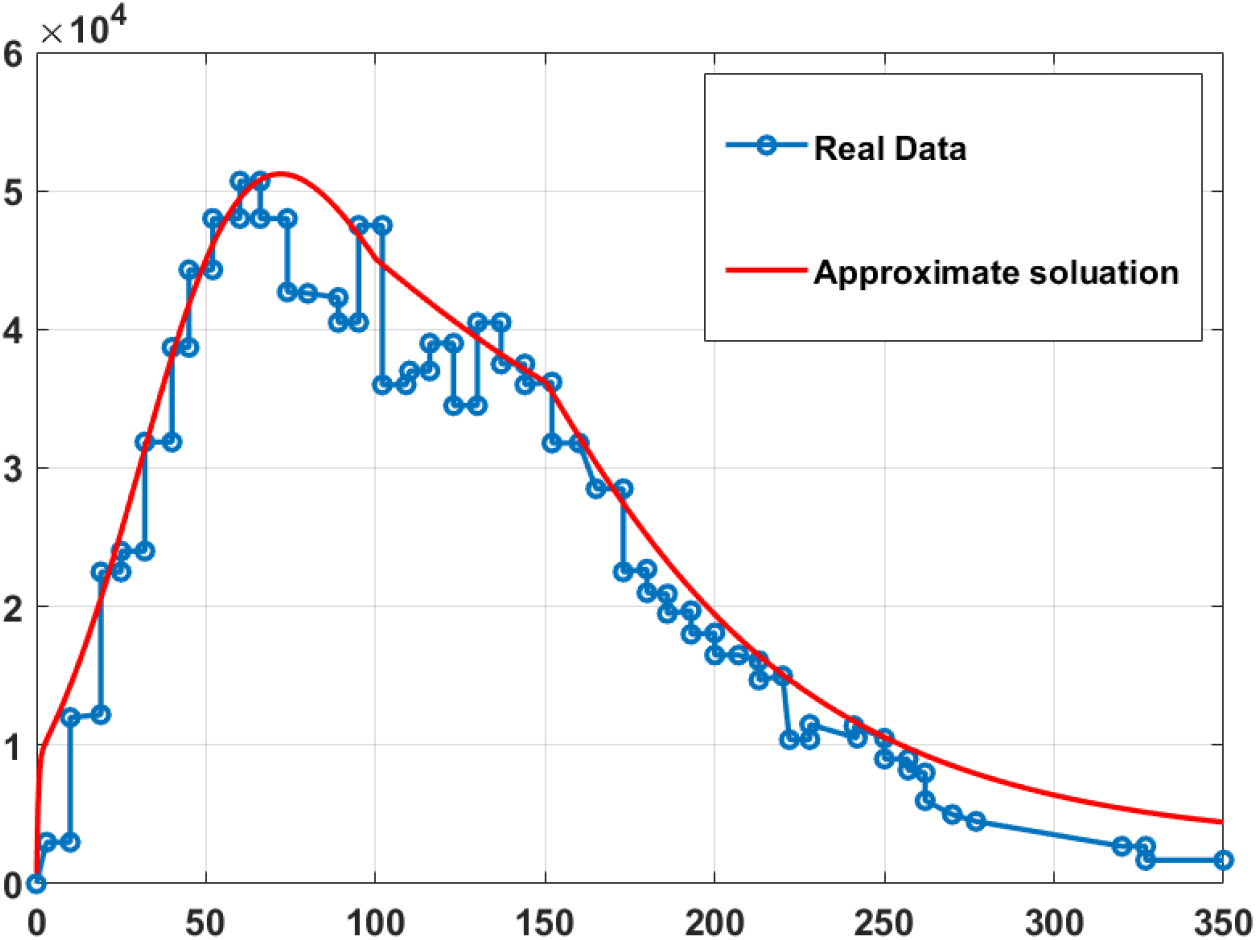
The numerical simulation for piecewise system of cholera (26)-(28) and (30)-(34) and without control-at *ξ* = 0.99, ν = 0.99 with Ψ(*t*) = 0.98*t* + 0.009, *ξ*(*t*) = −0.001*t* + 0.99, ν(*t*) = 1 − 0.001*t*, and 0 < *t* ≤ 100, 100 < *t* ≤ 150, 150 < *t* ≤ 350.

- For 0 < *t* ≤ 100, the sharp increase in cases is accurately captured by the approximate solution.
- In 100 < *t* ≤ 150, the peak is well-aligned, showing the model effectively predicts the maximum outbreak intensity.
- For *t* > 150, the decline is slower but consistent, reflecting a gradual reduction in cholera cases. Moreover, the simulation considers specific parameter variations (*ξ*(*t*), ν(*t*), Ψ(*t*)), and the agreement between the approximate solution and real data highlights the model’s ability to simulate disease dynamics under varying conditions.

Regarding the Biological significance:

- The rise and fall of cholera cases correspond to infection spread, peak outbreaks, and mitigation phases.
- This model can be utilized to study intervention strategies by adjusting parameters like *ξ*(*t*) and ν(*t*), representing external or control factors.

Figure 4’s simulation Ψ(*t*) = *t* provides a more accurate representation of cholera dynamics in environments where transmission follows natural epidemic curves. Figure 3 is useful for scenarios with accelerating factors in the early outbreak phases.

**Figure 4:**
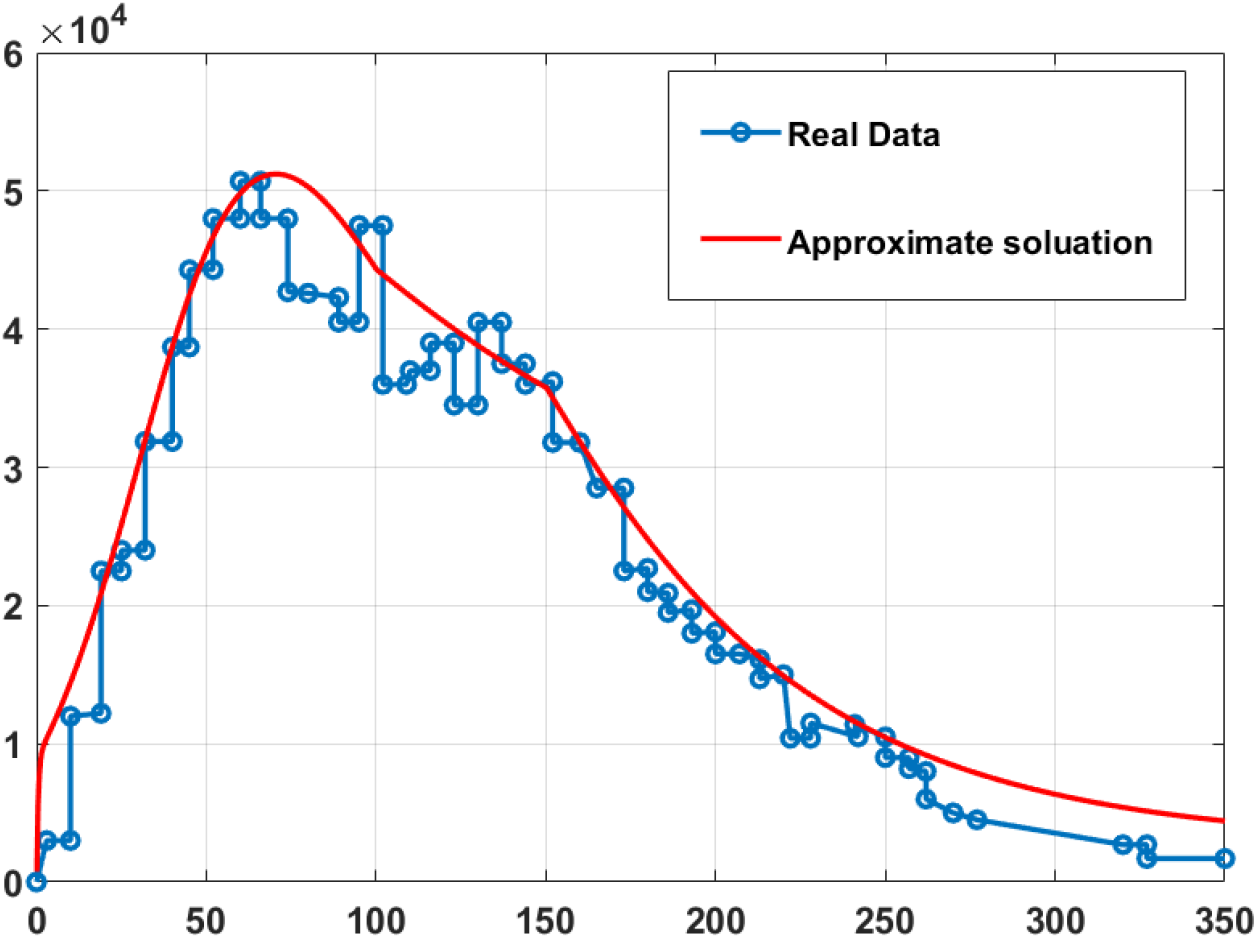
The numerical simulation for piecewise system cholera (26)-(28) and (30)-(34) and without control at *ξ* = 0.99, ν = 0.99 with Ψ(*t*) = *t, ξ*(*t*) = −0.001*t* + 0.99, ν(*t*) = −0.001*t* + 0.99, and 0 < *t* ≤ 100, 100 < *t* ≤ 150, 150 < *t* ≤ 350.

Figure 5 provides a visual representation of the impact of control measures on the dynamics of a cholera outbreak. Compared to those without control, the control curves demonstrate reductions in infected individuals (𝕀) and potentially environmental bacteria (𝔹). This indicates that the control measures are successfully mitigating cholera transmission. Additionally, the “with control” curve for susceptible individuals (𝕊) may exhibit a slower decline, suggesting increased protection for the population.

**Figure 5:**
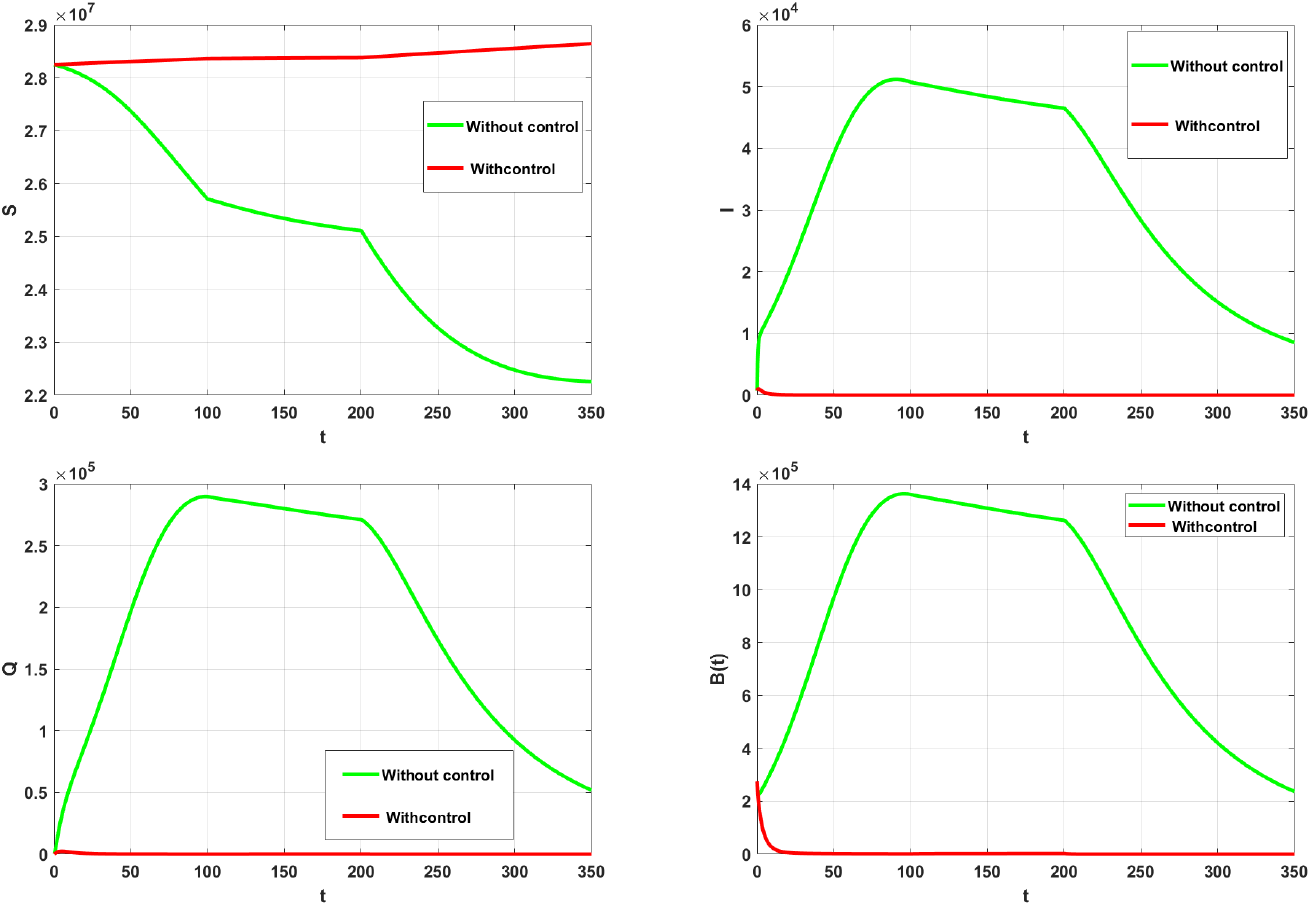
The numerical simulation for piecewise system cholera (26)-(28) and (30)-(34) with and without control- at *ξ* = 0.95, ν = 0.98 Ψ(*t*) = (0.98*t* + 0.97)^0.99^, *ξ*(*t*) = −0.001*t* + 0.90, ν(*t*) = −0.001*cos*^2^(*t/*10) + 0.95, and 0 < *t* ≤ 100, 100 < *t* ≤ 200, 200 < *t* ≤ 350.

Figure 6 demonstrates how varying the values of *ξ*, and ν (potentially related to control measures) can significantly impact cholera dynamics.

**Figure 6:**
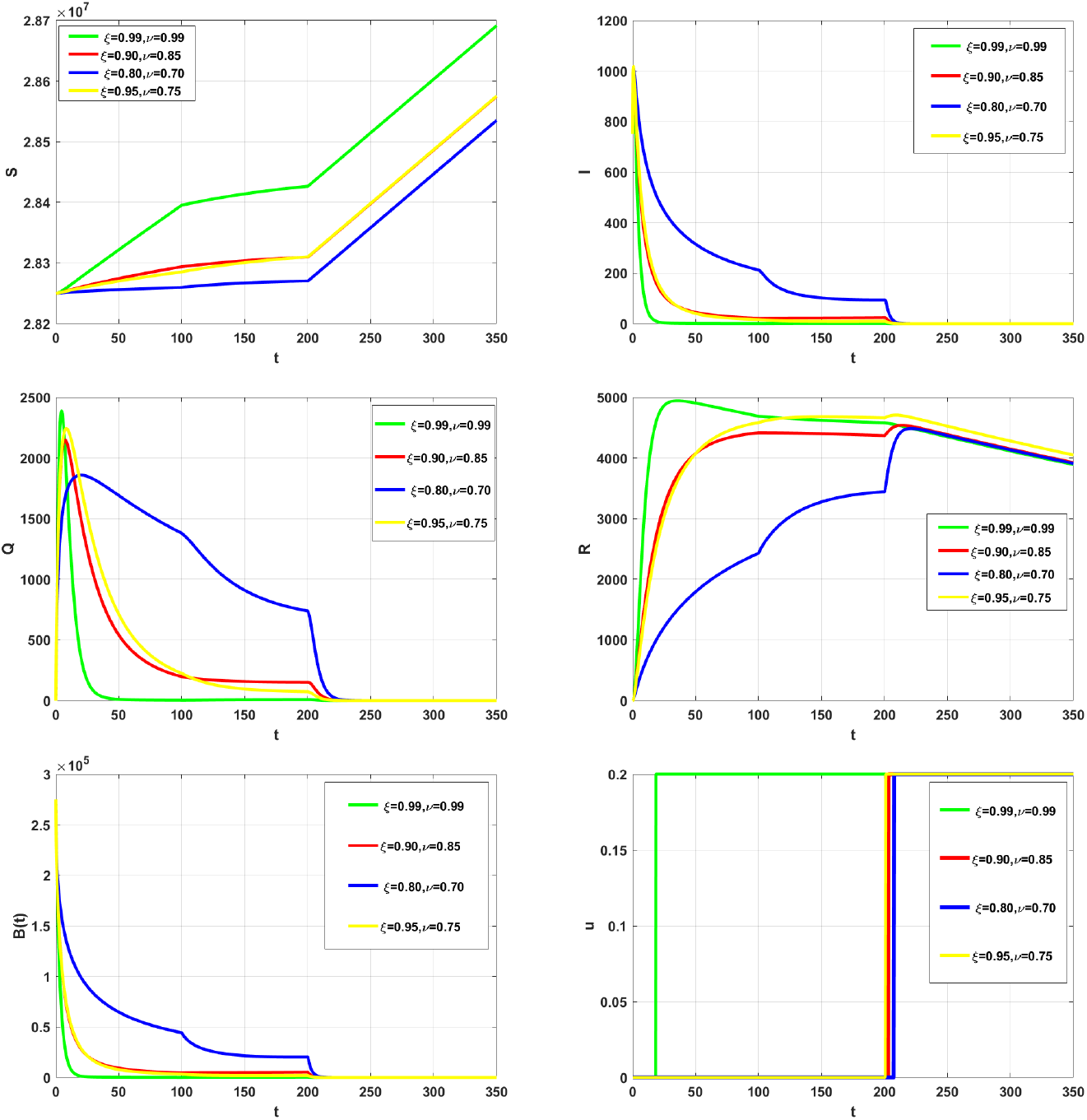
The numerical simulation for piecewise system of cholera (26)-(28) and (30)-(34) at Ψ(*t*) = (0.98*t* + 0.97)^0.99^, *ξ*(*t*) = −0.001*t* + 0.90, ν(*t*) = 0.95 − 0.001*cos*^2^(*t/*10), with different values of *ξ*, ν, and control case in and 0 < *t* ≤ 100, 100 < *t* ≤ 200, 200 < *t* ≤ 350.

Figure 7 effectively illustrates the impact of different values of *ξ*, ν, *ξ*(*t*), ν(*t*), and control strategies on the dynamics of a cholera outbreak through numerical simulations of a piecewise system. The graphs highlight key aspects of disease progression, including susceptible, infected, and recovered populations and environmental bacterial concentrations. The results demonstrate the effectiveness of timedependent interventions in reducing infection rates and bacterial loads, emphasizing the importance of adaptive control measures over different phases of the outbreak. This piecewise approach mirrors real-world public health responses, providing valuable insights into optimal strategies for mitigating cholera transmission and safeguarding public health.

**Figure 7:**
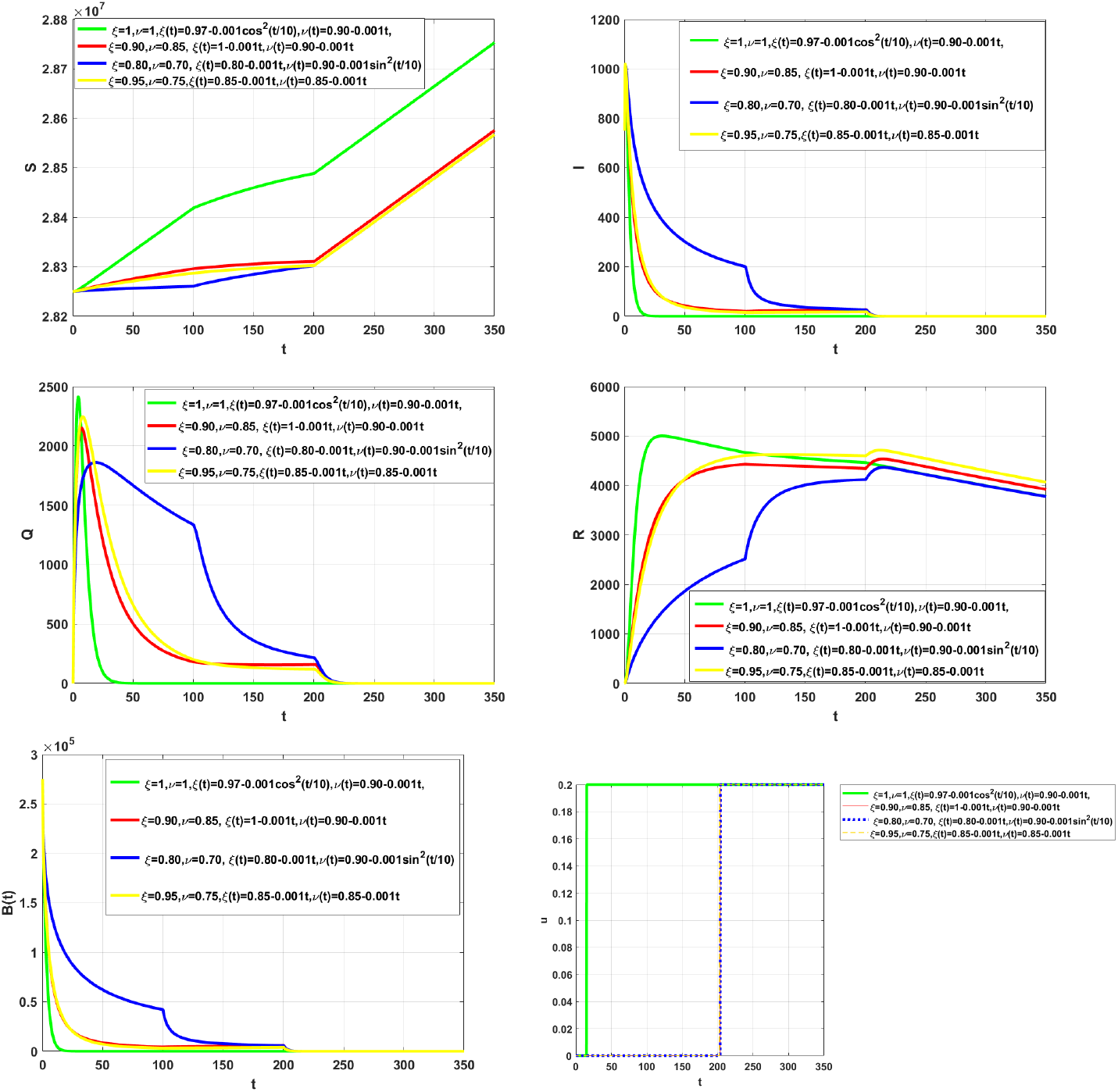
The numerical simulation for piecewise system of cholera (26)-(28) and (30)-(34) at Ψ(*t*) = (0.98*t* + 0.009) with varied values of *ξ*, ν, *ξ*(*t*), ν(*t*), and control case in and 0 < *t* ≤ 100, 100 < *t* ≤ 200, 200 < *t* ≤ 350.

Figure 8 presents numerical simulations for a piecewise cholera model, demonstrating how varying functions Ψ(*t*), *ξ*(*t*), ν(*t*) influence the system dynamics over time. The results compare scenarios with different forms of time-dependent interventions, highlighting their effects on susceptible, infected, and recovered populations, as well as bacterial concentration in the environment. The gradual decline in infection and bacterial load across different intervals suggests that control measures such as improved sanitation, vaccination, or treatment lead to reduced transmission and environmental contamination. The figure underscores the importance of dynamic, adaptive strategies in effectively managing cholera outbreaks over extended periods.

**Figure 8:**
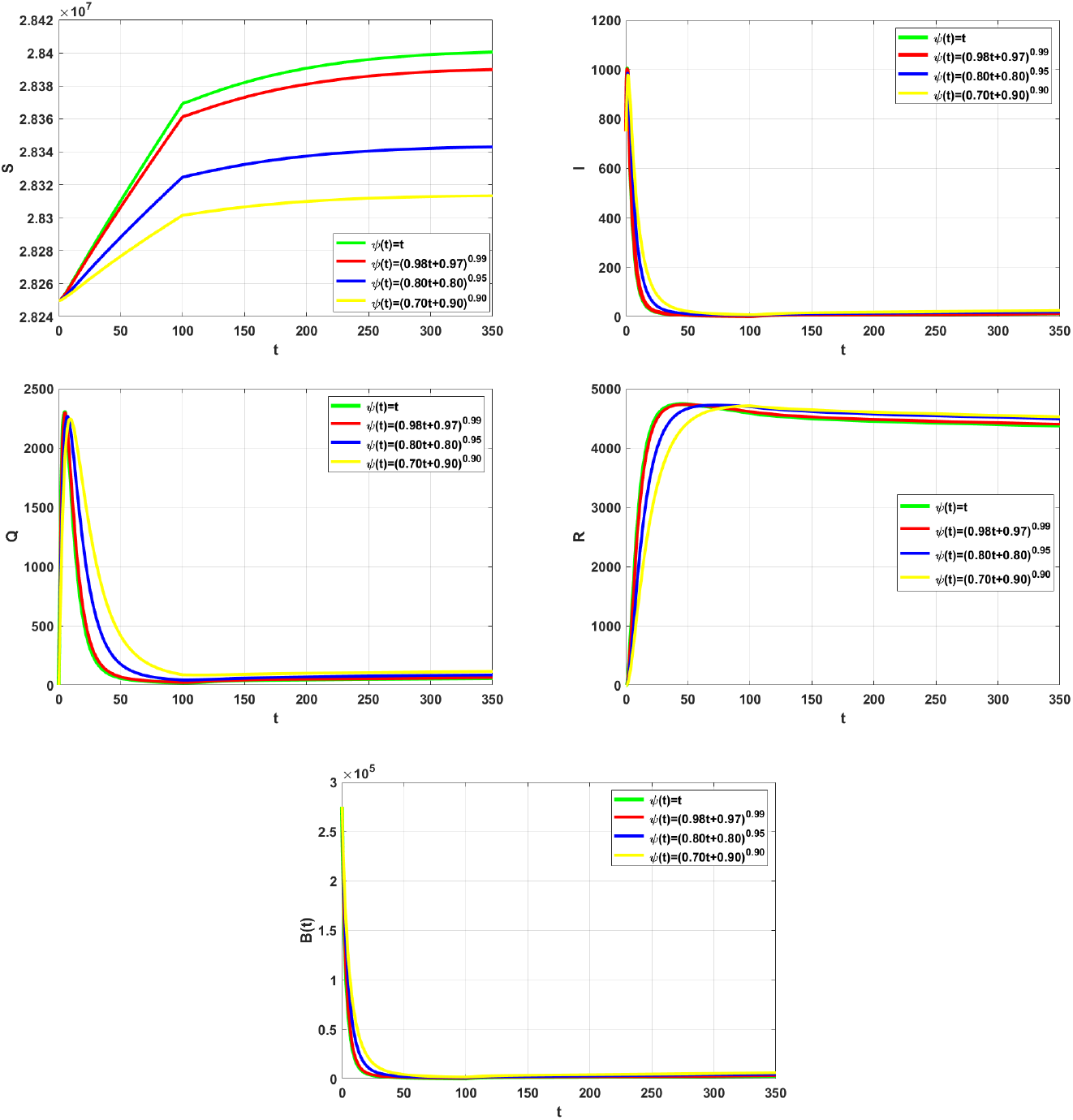
The numerical simulation for piecewise system of cholera (26)-(28) and (30)-(34)- at *ξ* = 0.95, ν = 0.98 with varied values Ψ(*t*), *ξ*(*t*) = −0.001*t* + 0.90, ν(*t*) = −0.001*cos*^2^(*t/*10) + 0.95, and control case in 0 < *t* ≤ 100, 100 < *t* ≤ 350.

Figure 9 showcases the results of a numerical simulation for a piecewise cholera model, illustrating the influence of varying functions Ψ(*t*), *ξ*(*t*), ν(*t*) over different time intervals in the control case. The graphs depict the progression of key variables, including susceptible, infected, recovered populations, and environmental bacterial concentration. The results emphasize the effectiveness of dynamic control measures, such as time-dependent vaccination, sanitation, and treatment interventions, in reducing infection rates and environmental contamination. The figure highlights the importance of sustained, adaptive strategies in mitigating the spread of cholera, demonstrating noticeable reductions in cases and bacterial load as control efforts intensify across the specified time segments.

**Figure 9:**
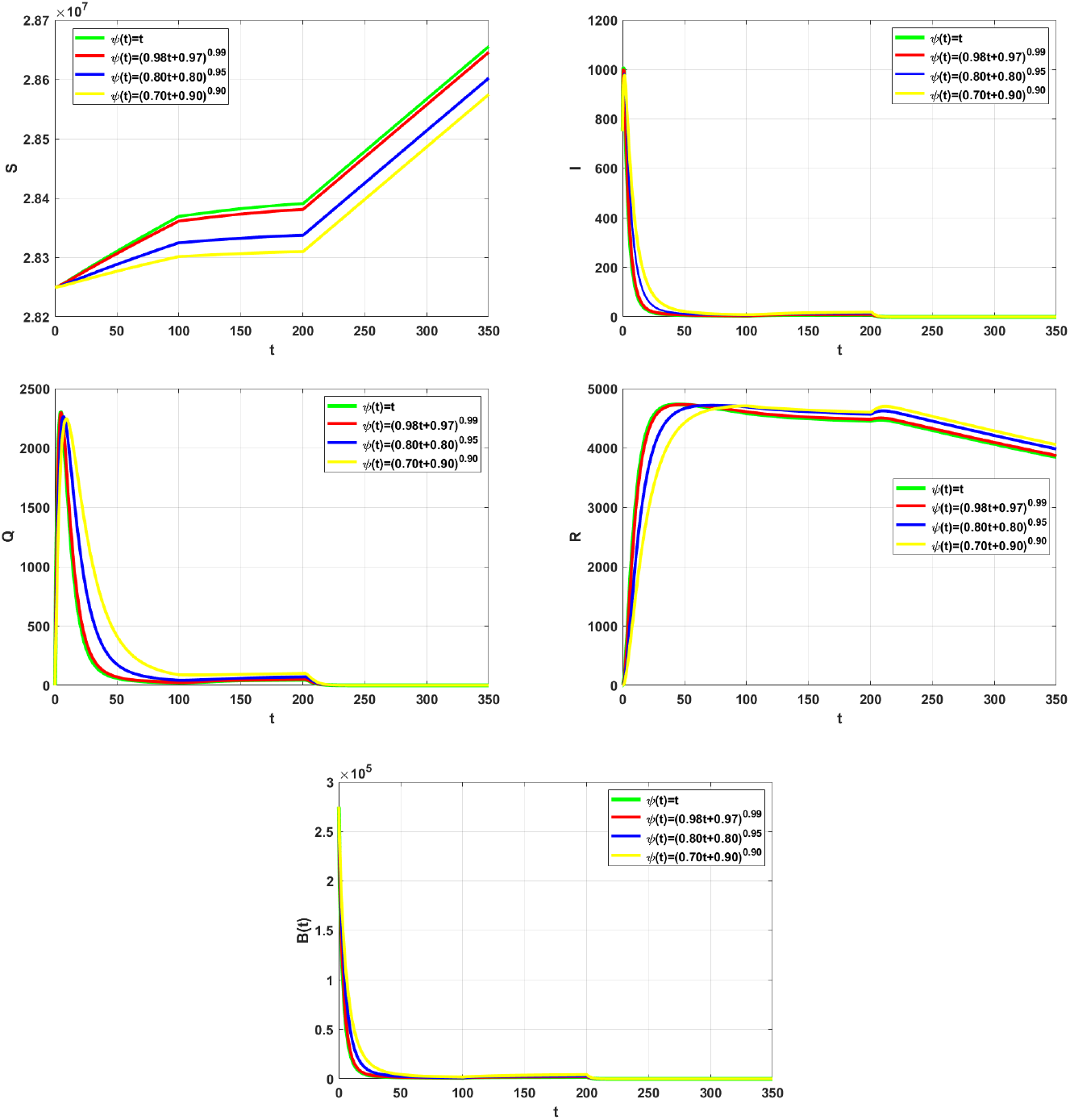
The numerical simulation for piecewise system of cholera (26)-(28) and (30)-(34)- at *ξ* = 0.95, ν = 0.98 with different Ψ(*t*), *ξ*(*t*) = −0.001*t* + 0.90, ν(*t*) = −0.001*cos*^2^(*t/*10) + 0.95, and control case in 0 < *t* ≤ 100, 100 < *t* ≤ 200, 200 < *t* ≤ 350.

**Figure 10:**
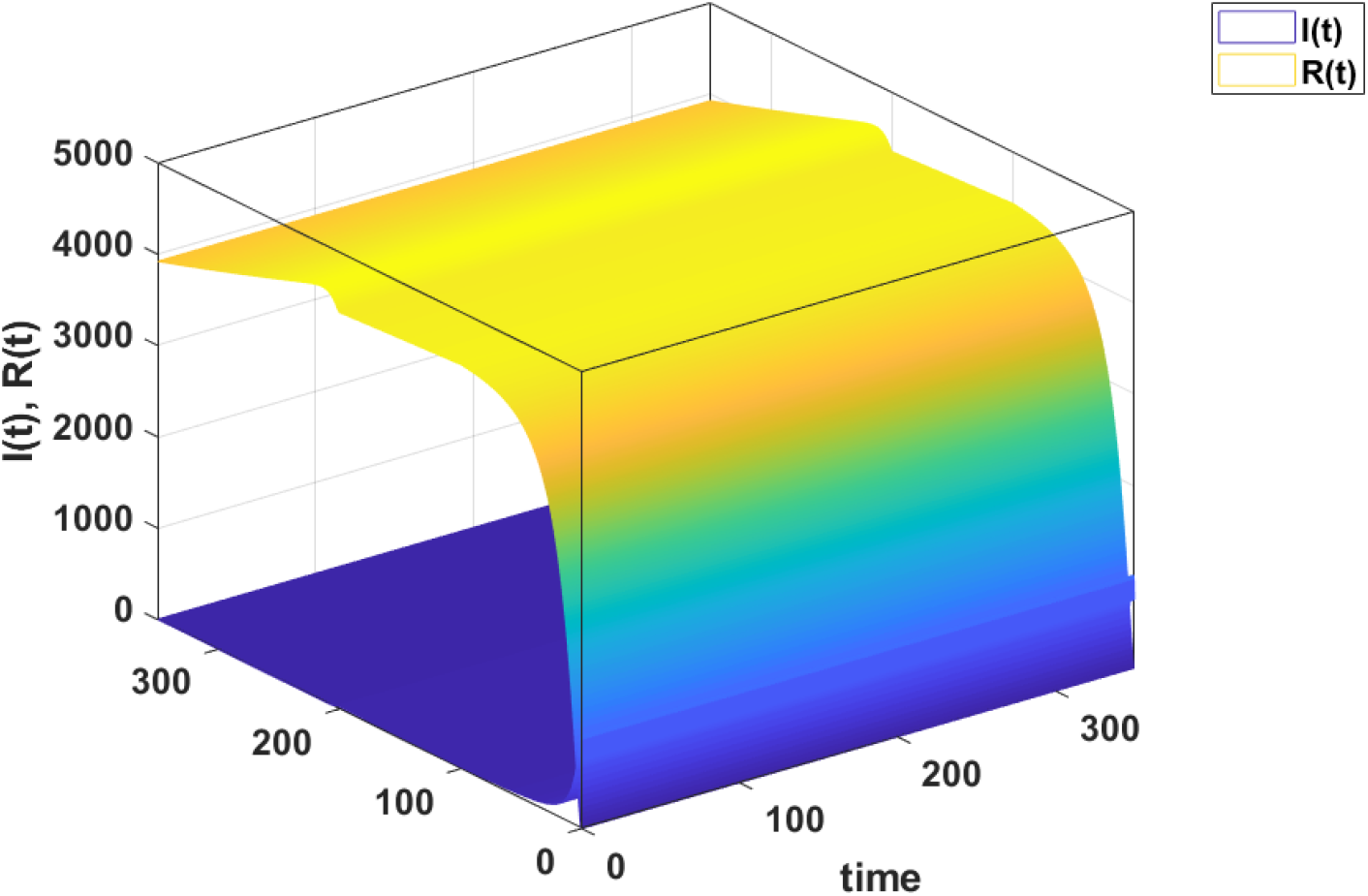
The numerical simulation for relationship between *R*(*t*), *I*(*t*) and control case- at *ξ* = 0.90, ν = 0.85, *ξ*(*t*) = −0.001*t* + 1, ν(*t*) = −0.001*t* + 0.90 with Ψ(*t*) = 0.98*t* + 0.009, and 0 < *t* ≤ 100, 100 < *t* ≤ 150, 150 < *t* ≤ 350.

This figure presents a 3D numerical simulation illustrating the relationship between the infected population 𝕀(*t*) and the recovered population ℝ(*t*) over time under different control scenarios. The smooth transition in the graph highlights the progression of the epidemic, showing the increase in infections initially, followed by a rise in recovered individuals as time advances. The influence of control measures such as Ψ(*t*), *ξ*(*t*), ν(*t*) is reflected in the gradual reduction of 𝕀(*t*) and the corresponding growth in ℝ(*t*). The visualization underscores the effectiveness of time-dependent interventions in mitigating the outbreak, leading to faster recovery and a lower peak infection rate. This 3D representation provides valuable insights into the dynamics of cholera progression and the importance of adaptive strategies to control its spread.

Table 2 presents a comparison of the objective functional *J* under different control strategies, reflecting the impact of varying Ψ(*t*) functions on the cholera model. The results highlight a significant reduction in *J* when control measures are applied, as evidenced by the consistently lower values in the “J with control” column compared to the “J without control” column. The most substantial improvement is observed with Ψ(*t*) = (0.97*t* + 0.80)^0.90^, suggesting that this control function yields the best outcome in minimizing the objective functional. This comparison underscores the effectiveness of time-dependent control strategies in reducing the severity of the outbreak, demonstrating the importance of adaptive interventions tailored to the dynamics of the epidemic.

**Table 2:** Comparing the objective functional value *J* when *ξ* = 0.85 ν = 0.90 *ξ*(*t*) = 0.97 − 0.001*cos*^2^(*t/*10) ν(*t*) = 1 − 0.008*t* and 0 < *t* ≤ 100, 100 < *t* ≤ 150, 150 < *t* ≤ 350.

| $\Psi(t)$ | J with control | J without control |
| --- | --- | --- |
| $\Psi(t) = t$ | $1.988 \times 10^5$ | $1.1767 \times 10^7$ |
| $\Psi(t) = (0.97t + 0.80)^{0.90}$ | $2.7774 \times 10^5$ | $1.0667 \times 10^7$ |
| $\Psi(t) = (0.80t + 0.007)$ | $2.4126 \times 10^5$ | $1.1196 \times 10^7$ |
| $\Psi(t) = (t^{0.98})$ | $2.1376 \times 10^5$ | $1.1545 \times 10^7$ |

## 7 Conclusion

This study introduced a novel crossover mathematical model for cholera that combines integer-order derivatives, fractal fractional-order derivatives, and Ψ-Caputo fractal variable-order fractional derivatives within a unified framework. By incorporating a non-standard kernel Ψ(*t*), the model effectively captures the dynamics of cholera across three distinct time intervals, allowing for greater adaptability and accuracy in representing real-world outbreaks.

The stability analysis confirmed the robustness of the model’s steady states, and the comparative analysis with real data from the 2017-2018 cholera outbreak in Yemen validated the model’s predictive capability. The formulation and implementation of an optimal control problem further demonstrated the model’s potential in mitigating disease spread. The use of the discretized Ψ-finite difference method provided an efficient and reliable numerical approach for solving the crossover optimality system.

Simulation results highlighted the superiority of the crossover-controlled system in reducing the prevalence of cholera, underscoring its effectiveness as a disease management tool. This work contributes to the growing body of research on fractional and fractal-order models, offering a powerful framework for addressing complex epidemiological challenges.

Future research may explore the extension of this model to other infectious diseases, incorporate additional real-world datasets, and refine the control strategies to enhance public health interventions further.

## Data Availability

All relevant data are within the manuscript and its Supporting Information files.

## References

[1] World Health Organization, 2022,https://www.who.int/news/item/20-05-2022-world-health-statistics-2022

[2] M. Ali, A. R. Nelson, A. L. Lopez, & D. A. Sack, The global burden of cholera. Bulletin of the World Health Organization, 93(3), 209–218. 10.2471/BLT.14.150109

[3] C. T. Codeco, Endemic and epidemic dynamics of cholera: The role of the aquatic reservoir. BMC Infectious Diseases, 1(3), 1471–2334, (2001), doi:10.1186/1471-2334-1-1.

[4] A. P. Lemos-Paião, C. J. Silva1, D.F. M. Torres, E. Venturino, (2020). Optimal Control of Aquatic Diseases: A Case Study of Yemen’s Cholera Outbreak, Journal of Optimization Theory and Applications 185:1008–1030

[5] Y. He, Z. Wang, Stability Analysis and Optimal Control of a Fractional Cholera Epidemic Model. Fractal and Fractional. 6(3),157, (2022). 10.3390/fractalfract6030157

[6] A. Huppert, and G. Katriel, (2013). Mathematical modelling and prediction in infectious disease epidemiology. Clinical Microbiology and Infection, 19(11), 999–1005.

[7] S. G. Samko, A. A. Kilbas, and O. I. Marichev, Fractional integrals and derivatives, Gordon and Breach Science Publishers, Yverdon, (1993).

[8] A. A. Kilbas, H. M. Srivastava, and J. J. Trujillo, Theory and Applications of Fractional Differential Equations, North-Holland Math. Stud.,Elsevier Science B.V., Amsterdam, (2006).

[9] R. Almeida, A. B. Malinowska, and M. T. T. Monteiro, Fractional differential equations with a Caputo derivative with respect to a kernel function and their applications, Math. Methods Appl. Sci., 41 (1), 336–352, (2018).

[10] J. E. Solís-Pérez1, J.F. Gómez-Aguilar, Variable-order fractal-fractional time delay equations with power, exponential and Mittag-Lefer laws and their numerical solutions, Engineering with Computers, 38,555–577, (2022)

[11] International Medical Corps UK: Emergency treatment and prevention of cholera in Yemen (2018). https://www.internationalmedicalcorps.org.uk/emergency-treatment-and-preventioncholera-yemen. Accessed 25 May 2018

[12] O. P. Agrawal, A general formulation and solution scheme for fractional optimal control problems, Nonlinear Dynam., 38, 1, 323–337, 2004

[13] N. H. Sweilam and S. M. Al-Mekhlafi and A. O. Albalawi and J. A. T. Machado, Optimal control of variable-order fractional model for delay cancer treatments, Appl. Math. Model., 89, 1557–1574, 2021

[14] N. Sweilam and F. Rihan and S. M. Al-Mekhlafi A fractional-order delay differential model with optimal control for cancer treatment based on synergy between anti-angiogenic and immune cell therapies, Discrete Contin Dyn Syst Ser., 13, 9, 2403–2424, 2020,

[15] R. E. Mickens, Nonstandard finite difference models of differential equations, World Scientific, Singapore, (2005).

[16] A. Atangana, S. Qureshi, Modeling attractors of chaotic dynamical systems with fractal-fractional operators, Chaos, Solitons and Fractals, 123 (2019) 320–337.

[17] N.H. Sweilam, S.M. AL-Mekhlafi, D.G. Mohamed, Novel chaotic systems with fractional differential operators: Numerical approaches, Chaos, Solitons and Fractals, (2020), 10.1016/j.chaos.2020.110475

[18] K. Diethelm, The Analysis of Fractional Differential Equations: An Application-Oriented Exposition Using Differential Operators of Caputo Type. Springer, Berlin, (2010).

[19] R. T. Alqahtani, B. Ahmad, D. Baleanu, On variable-order Ψ-Caputo fractional differential equations with applications. Mathematics, 9(2), 156, (2021).

[20] R. L. Neilan, S. Lenhart, An Introduction to Optimal Control with an Application in Disease Modeling. American Mathematical Society, (2010).

[21] N. H. Sweilam, S. M. AL-Mekhlafi, W.S. Abdel Kareem, G. Alqurishi, Comparative Study of Crossover Mathematical Model of Breast Cancer Based on Ψ-Caputo Derivative and Mittag-Leffler Laws: Numerical Treatments, Symmetry, 16(9), 1172, (2024).

[22] N. H. Sweilam, S. M. AL-Mekhlafi, W.S. Abdel Kareem, G. Alqurishi, A New Crossover Dynamics Mathematical Model of Monkeypox Disease Based on Fractional Differential Equations and the Ψ-Caputo Derivative: Numerical Treatments, Alexandria Engineering Journal, 111, 181–193, (2025).

[23] F. Abbas, A. Ghaffar, A. Akgul, A. Ahmad, G. Mustafa, A. S. Hendy, S. Ali O. Abdallah, N. S. Abd El-Gawaad, Analysis and dynamics of cholera epidemic system in society via fractal-fractional operator, Fractals, (2023), doi:10.1142/S0218348X24400528

[24] I. Podlubny, Fractional Differential Equations, Academic Press, New York, (1999).

[25] World Health Organization: Yemen: Weekly cholera bulletins

[26] Van denDriessche, P., Watmough, J., (2008). Further Notes on the Basic Reproduction Number. Berlin; Heidelberg: Springer Berlin Heidelberg.

[27] Van den Driessche P., (2002). Watmough J. Reproduction numbers and sub-threshold endemicequilibria for compartmental models of disease transmission. Math Biosci. 180:29–48. doi: 10.1016/S0025-5564(02)00108-6

[28] Allen, L. J., (2007). Introduction to Mathematical Biology. Pearson; Prentice Hall.

